# Evaluation of Corneal Subbasal Nerve Plexus Alterations in ARSACS and SPG7 by In Vivo Corneal Confocal Microscopy

**DOI:** 10.64898/2026.06.22.26356257

**Authors:** Umit Yasar Guleser, Nazan Akkaya, Cem Kesim, Özgür Öztop Çakmak, Melisa Zisan Karslioglu, Ayşe Nazlı Başak, Sibel Ertan, Murat Hasanreisoglu, Atay Vural

## Abstract

**Purpose:** To investigate corneal subbasal nerve plexus alterations using in vivo corneal confocal microscopy (IVCM) in patients with Autosomal Recessive Spastic Ataxia of Charlevoix–Saguenay (ARSACS) and Spastic Paraplegia Type 7 (SPG7).

**Methods:** This cross-sectional pilot study included eight ARSACS patients, five SPG7 patients, and twenty age- and sex-matched healthy controls. All participants underwent neurological and ophthalmological examination followed by central corneal imaging using IVCM. Quantitative corneal nerve parameters were analyzed with automated software, and correlations with clinical severity scales were assessed.

**Results:** The mean age was 34.2 ± 3.4 years in controls, 34.5 ± 0.7 years in the ARSACS group, and 38.2 ± 3.5 years in the SPG7 group. Corneal nerve branch density (CNBD) and corneal nerve total branch density (CTBD) were significantly lower in ARSACS and SPG7 patients compared with healthy controls. CNFD, CNFL, CNFA, CNFW, and CNFrD were lower in ARSACS and SPG7 patients compared with healthy controls; however, these differences did not reach statistical significance. No statistically significant differences in IVCM parameters were detected between ARSACS and SPG7 patients. Spearman correlation analysis did not show significant correlations between corneal nerve parameters and FARS, SARA, ADL scores, or disease duration.

**Conclusion:** IVCM revealed reduced corneal nerve branching parameters in patients with ARSACS and SPG7. These findings indicate involvement of the corneal subbasal nerve plexus and support the potential role of corneal confocal microscopy as a non-invasive ocular imaging modality for evaluating peripheral neural alterations in hereditary spastic ataxias.

## Introduction

Hereditary ataxias constitute a heterogeneous group of neurodegenerative disorders characterized by progressive cerebellar dysfunction, frequently accompanied by pyramidal signs and peripheral neuropathy [1]. Within this spectrum, Autosomal Recessive Spastic Ataxia of Charlevoix–Saguenay (ARSACS) and Spastic Paraplegia Type 7 (SPG7) represent paradigmatic forms of spastic ataxia, sharing overlapping clinical features despite distinct genetic and pathophysiological mechanisms [2,3]. ARSACS typically presents in early childhood with cerebellar ataxia, lower-limb spasticity, and prominent sensorimotor peripheral neuropathy [4], whereas SPG7 usually manifests later in life and is characterized by progressive spastic paraplegia with variable cerebellar involvement [5]. This phenotypic overlap often complicates clinical stratification and underscores the need for objective biomarkers capturing both central and peripheral nervous system involvement.

Ocular involvement is increasingly recognized as a relevant feature in hereditary ataxias, with ARSACS and SPG7 exhibiting distinct yet partially overlapping ophthalmic findings. ARSACS is associated with retinal nerve fiber layer thickening, papillomacular folds, and foveal hypoplasia [6], while SPG7 more commonly presents with optic neuropathy, ptosis, or nystagmus [7]. Although retinal and optic nerve alterations have been extensively studied, the cornea—despite containing the highest density of sensory nerve fibers in the body—remains largely unexplored. Given that peripheral neuropathy is a hallmark of ARSACS and has also been reported in SPG7 [8,9], corneal subbasal nerve plexus alterations may represent an underrecognized marker of systemic neurodegeneration in these disorders.

In vivo corneal confocal microscopy (IVCM) enables non-invasive, high-resolution visualization and quantitative assessment of the corneal subbasal nerve plexus and has emerged as a sensitive biomarker of small fiber neuropathy in various systemic and neurodegenerative diseases [10]. While IVCM has demonstrated significant corneal nerve loss in Friedreich ataxia [11], corneal nerve involvement in ARSACS and SPG7 has not yet been investigated. We therefore hypothesized that corneal nerve alterations may constitute a shared but overlooked feature of these spastic ataxias. In this pilot study, we aimed to characterize corneal subbasal nerve plexus changes using IVCM in patients with ARSACS and SPG7 and to explore their relationship with clinical disease severity.

## Material and Methods

### Ethical Approval and Participant Consent

This study was approved by the local ethics committee of Koç University (Protocol number: 2024.344.IRB2.143). All procedures performed in this study adhered to the tenets of the Declaration of Helsinki. Written and verbal informed consent was obtained from all participants.

### Subjects

Eight patients diagnosed with ARSACS, five patients diagnosed with SPG7, and age and sex-matched 20 healthy controls who presented to the Department of Neurology at Koç University Hospital between 2021 and 2024, were included in the study. Each participant underwent detailed neurological examination, and investigations including blood for genomic analysis [12,13] (Table S1). Demographic data, including age, gender and disease duration were recorded. The Friedreich’s Ataxia Rating Scale (FARS) [14], the Scale for the Assessment and Rating of Ataxia (SARA) [15] and The Activities of Daily Living (ADL) Scale [16] of all patients were measured.

All subjects underwent full ophthalmological examination including autorefractometry, best corrected visual acuity (BCVA), slit-lamp biomicroscopy, non-contact tonometry and dilated fundoscopy and IVCM. Patients with any other autoimmune or neurological disease, diabetes, glaucoma, ocular surface disorders, contact lens use, history of ocular surgery, dry eye syndrome, meibomian gland dysfunction, allergic ocular surface disease, pterygium, corneal dystrophy, uveitis, or any additional condition that could affect ocular surface parameters and patients who cannot perform IVCM due to nystagmus were excluded from the study.

**Table S1.**
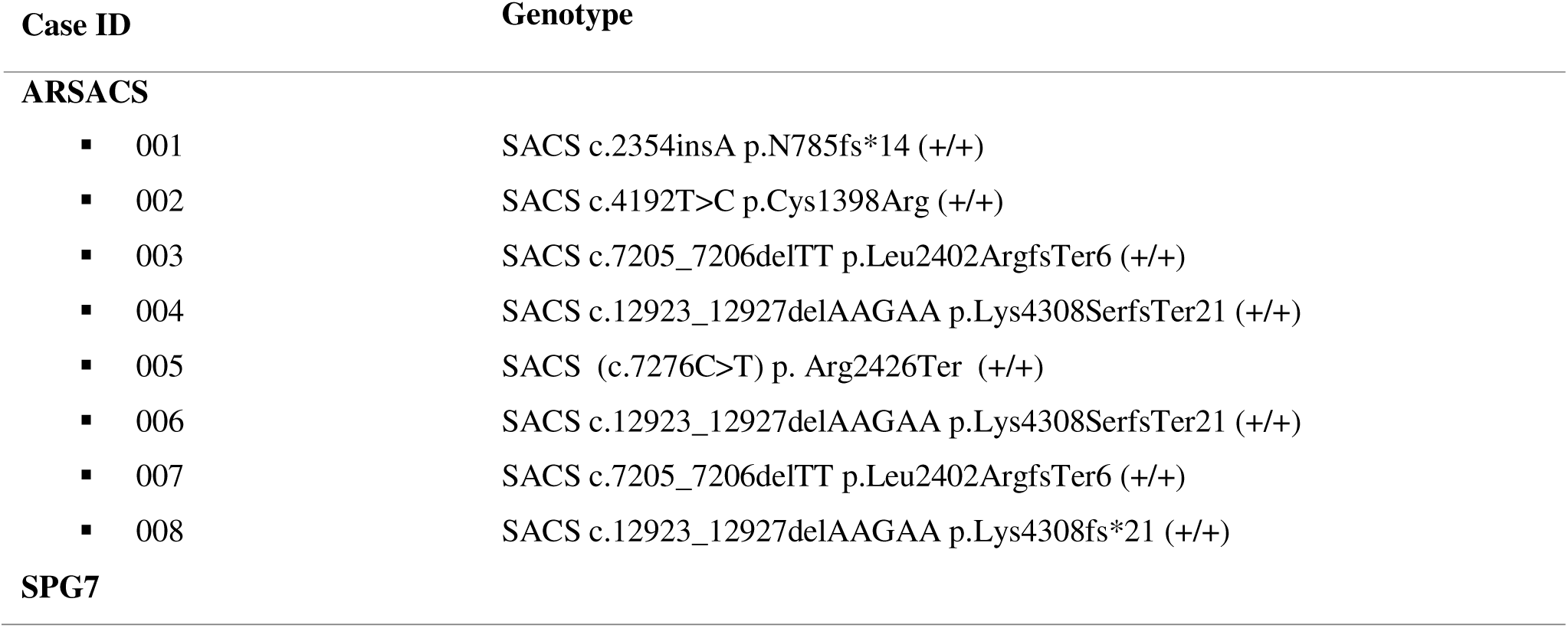
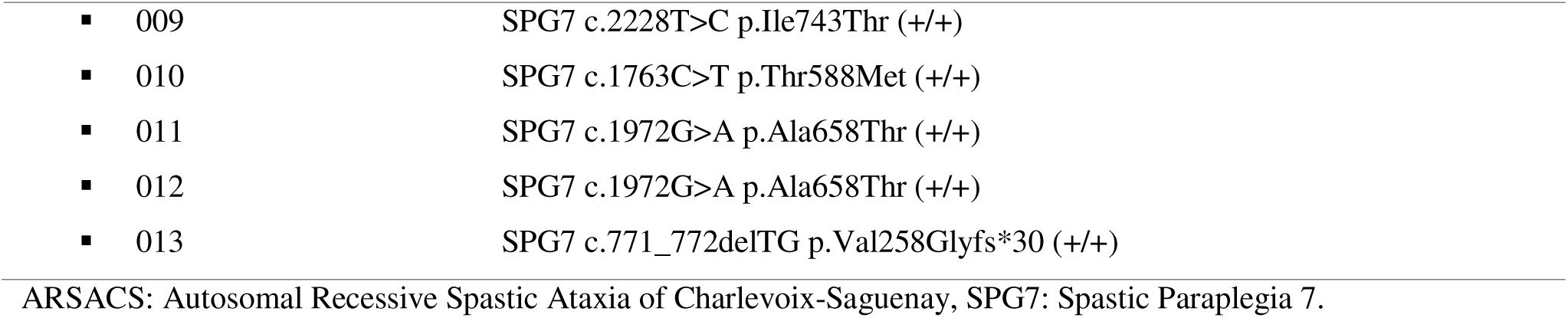
Genotypic distribution and identified pathogenic variants in ARSACS and SPG7 patients undergoing corneal confocal microscopy analysis.

| Case ID | Genotype |
| --- | --- |
| **ARSACS** |  |
| ▪ 001 | SACS c.2354insA p.N785fs*14 (+/+) |
| ▪ 002 | SACS c.4192T>C p.Cys1398Arg (+/+) |
| ▪ 003 | SACS c.7205_7206delTT p.Leu2402ArgfsTer6 (+/+) |
| ▪ 004 | SACS c.12923_12927delAAGAA p.Lys4308SerfsTer21 (+/+) |
| ▪ 005 | SACS (c.7276C>T) p. Arg2426Ter (+/+) |
| ▪ 006 | SACS c.12923_12927delAAGAA p.Lys4308SerfsTer21 (+/+) |
| ▪ 007 | SACS c.7205_7206delTT p.Leu2402ArgfsTer6 (+/+) |
| ▪ 008 | SACS c.12923_12927delAAGAA p.Lys4308fs*21 (+/+) |
| **SPG7** |  |
| ▪ 009 | SPG7 c.2228T>C p.Ile743Thr (+/+) |
| ▪ 010 | SPG7 c.1763C>T p.Thr588Met (+/+) |
| ▪ 011 | SPG7 c.1972G>A p.Ala658Thr (+/+) |
| ▪ 012 | SPG7 c.1972G>A p.Ala658Thr (+/+) |
| ▪ 013 | SPG7 c.771_772delTG p.Val258Glyfs*30 (+/+) |

ARSACS: Autosomal Recessive Spastic Ataxia of Charlevoix-Saguenay, SPG7: Spastic Paraplegia 7.

### Corneal Confocal Microscopy

The central corneal regions of all participants were scanned using corneal confocal microscopy with the Heidelberg Retinal Tomography 3 Rostock Cornea Module (Heidelberg Engineering GmbH, Heidelberg, Germany). Prior to image acquisition, topical anesthetic drops containing oxybuprocaine hydrochloride (4.0 g) and a carbomer-based lubricating gel (2 mg/g) were instilled in both eyes. Bilateral image acquisition was performed for five minutes by operators experienced in corneal confocal microscopy (UYG, CK, MZK). Following the imaging procedure, an experienced ophthalmologist (UYG) selected three high-quality images of the central corneal subbasal nerve plexus from each eye. Bilateral corneal imaging was performed in all participants. To avoid inter-eye correlation and reduce statistical bias, mean values obtained from both eyes were used for statistical analysis. The selected images, with a field of view of 400 × 400 µm², were analyzed using ACCMetrics software (Imaging Science and Biomedical Engineering, Manchester, UK), a fully automated image analysis program incorporating machine-learning–based algorithms [17–20] (Figure 1). Quantitative parameters obtained from the analysis included corneal nerve fiber density (CNFD), corneal nerve branch density (CNBD), corneal nerve fiber length (CNFL), corneal nerve total branch density (CTBD), corneal nerve fiber area (CNFA), corneal nerve fiber width (CNFW), and corneal nerve fractal dimension (CNFrD).

**Fig. 1.**
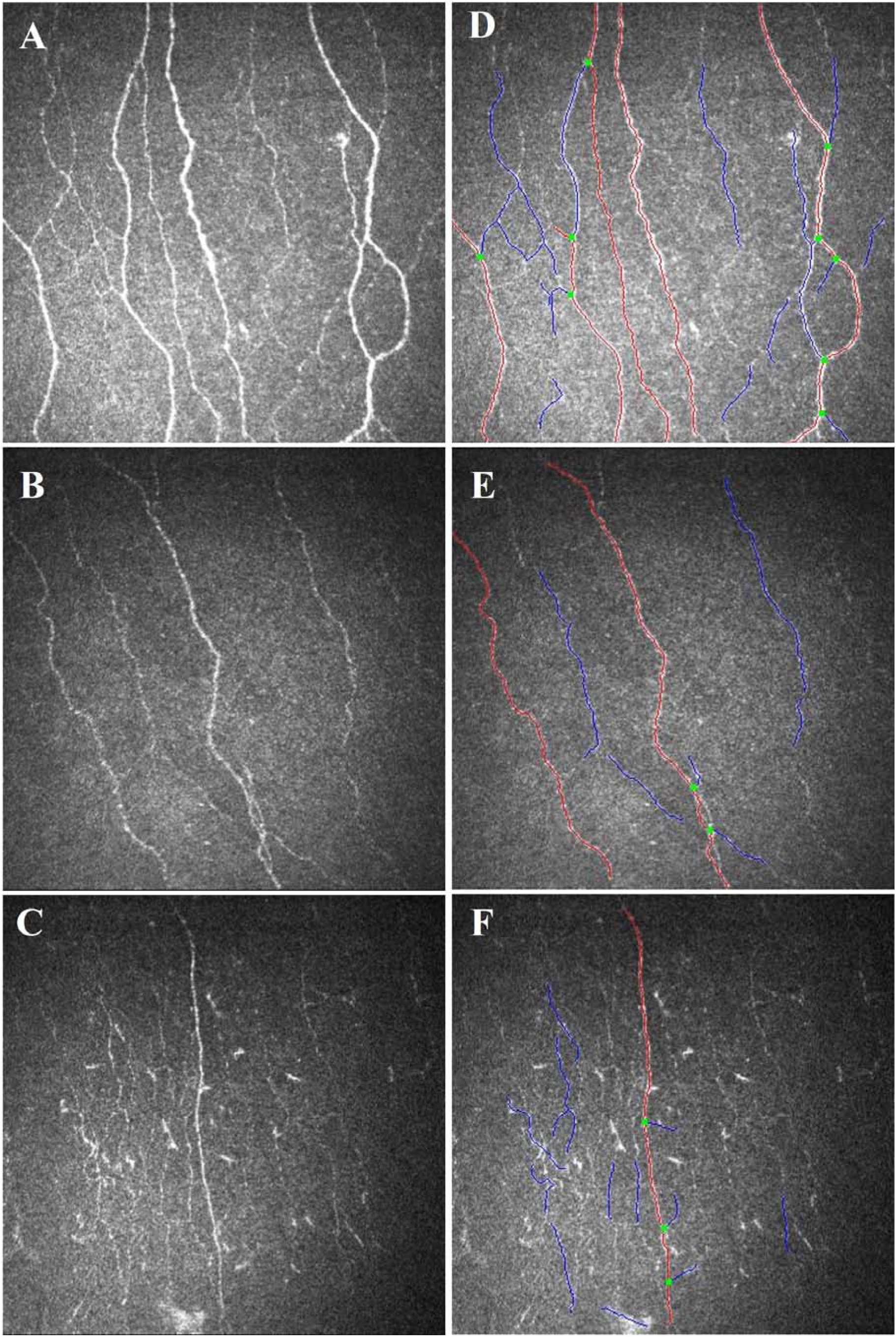
In vivo corneal confocal microscopy (IVCM) images of a healthy control, an Autosomal Recessive Spastic Ataxia of Charlevoix-Saguenay (ARSACS) patient and a Spastic Paraplegia 7 (SPG7) case. Main nerve fibers are automatically identified and highlighted in red, branches in blue, and branch points in green. A. IVCM image of a healthy control subject. B. Image of ARSACS patient with identifiable corneal subbasal nerve loss compared to control. C. IVCM image of SPG 7 patient with corneal subbasal nerve loss compared with control. D. Corresponding image of the control (A) analyzed by ACCMetrics Software (CNFD = 31.2480, CNBD = 56.2464, CNFL = 18.0333, CTBD = 74.9952, CNFA = 0,0079, CNFW = 0.0223, CFracDim = 1.5068). E. Corresponding image of ARSACS patient (B) analyzed by ACCMetrics Software (CNFD = 12.4992, CNBD = 12.4992, CNFL = 8.0922, CTBD = 12.4992 CNFA = 0.0024, CNFW = 0.0216, CFracDim = 1.4168). F. Corresponding image of SPG 7 patient (C) analyzed by ACCMetrics Software (CNFD = 6.2496, CNBD = 18.7488, CNFL = 6.6925, CTBD = 24.9984 CNFA = 0.0045, CNFW = 0.0199, CFracDim = 1.3636). CNFD, corneal nerve fiber density; CNBD, corneal nerve branch density; CNFL, corneal nerve fiber length; CTBD, corneal nerve total branch density; CNFA, corneal nerve fiber area; CNFW, corneal nerve fiber width; CNFrD, corneal nerve fractal dimension.

### Statistical Analysis

Statistical analyses were performed using SPSS Statistics software, version 25 (IBM Corp., Armonk, NY, USA). Descriptive statistics were reported as mean ± standard deviation, median, and interquartile range (25th–75th percentiles), as appropriate according to data distribution. The normality of continuous variables was assessed using the Shapiro–Wilk test, and visual inspection of Q–Q plots. Baseline demographic and clinical characteristics were compared among healthy controls, ARSACS patients, and SPG7 patients. Categorical variables were compared using the Chi-square test or Fisher’s exact test when expected cell counts were small. When significant overall group differences were identified, pairwise post hoc comparisons were performed using Dunn’s test with Bonferroni correction for multiple comparisons. For comparisons of continuous variables between two groups, the Mann–Whitney *U* test was used. Associations between clinical scale scores and corneal confocal microscopy parameters were assessed using Spearman’s rank correlation coefficient (ρ). A two-sided p-value of less than 0.05 was considered statistically significant.

## Results

Demographic and clinical characteristics of the study population are summarized in Table 1. There were no statistically significant differences among the groups with respect to age or sex distribution. The mean age was 34.2 ± 3.4 years in the control group, 34.5 ± 0.7 years in the ARSACS group, and 38.2 ± 3.5 years in the SPG7 group (p = 0.81). Sex distribution was comparable across groups (p = 0.70). The mean disease duration was similar between the ARSACS and SPG7 groups, measuring 27.5 ± 6.3 years and 27.6 ± 3.7 years, respectively (p = 0.82). Clinical severity scores did not differ significantly between patient groups. The mean FARS score was 2.93 ± 0.86 in the ARSACS group and 2.7 ± 0.43 in the SPG7 group (p = 0.65). Likewise, SARA scores were comparable between ARSACS (14.37 ± 3.84) and SPG7 patients (11.2 ± 2.14; p = 0.37). ADL scale scores also showed no significant difference between the two patient groups (11.06 ± 2.83 vs. 12.1 ± 1.84, p = 0.61).

**Table 1.**
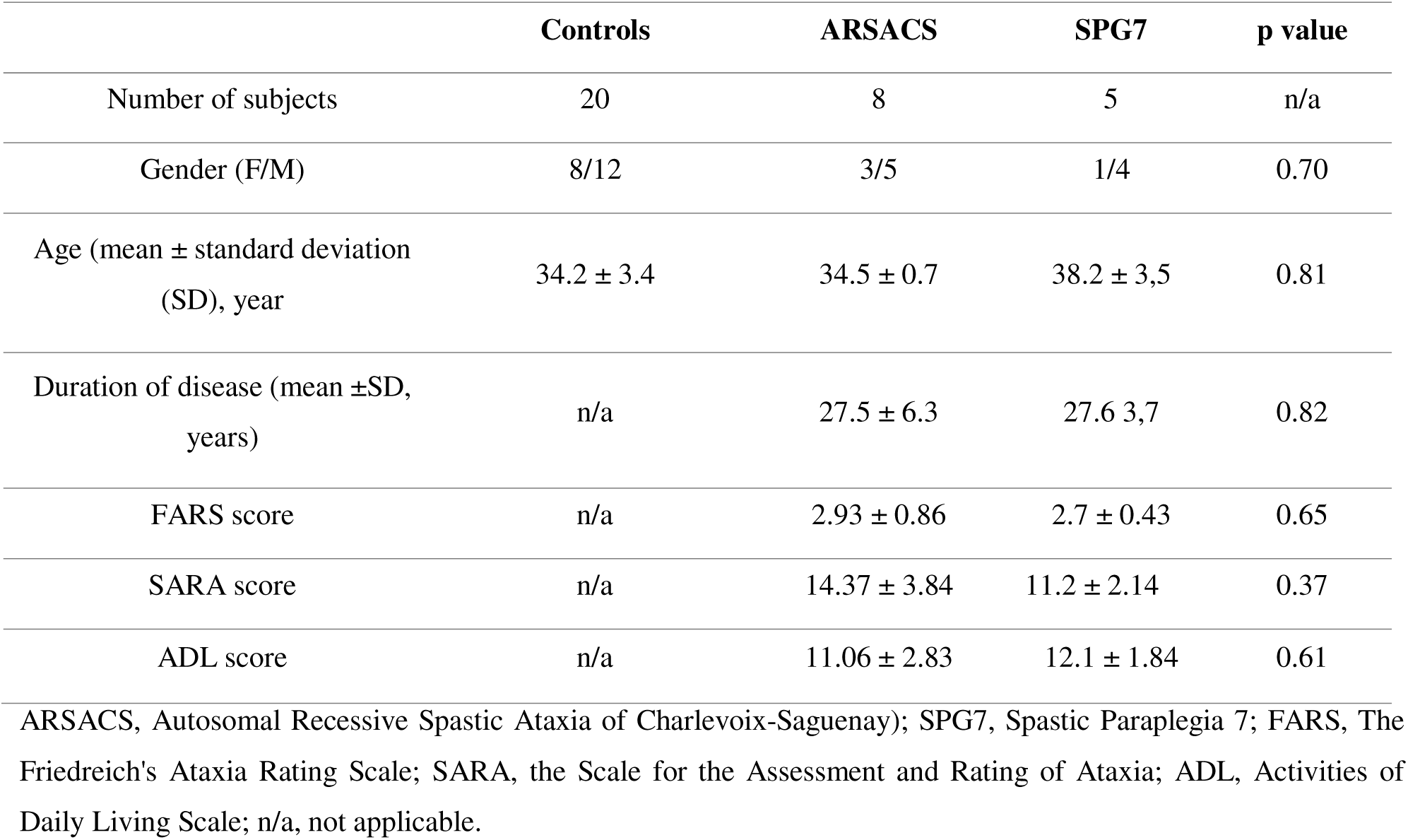
Demographic and clinical characteristics of the study population.

Corneal confocal microscopy parameters are summarized in Table 2. Compared with healthy controls, both ARSACS and SPG7 patients showed lower values across all IVCM parameters. CNBD and CTBD were significantly lower in ARSACS patients than controls (p = 0.045 and p = 0.042, respectively), and similar reductions were observed in SPG7 patients (p = 0.01 and p = 0.025, respectively). Although the remaining IVCM parameters also showed a trend toward lower values in both patient groups, these differences did not reach statistical significance. No significant differences were detected between ARSACS and SPG7 groups.

**Table 2.** Comparison of corneal nerve parameters assessed by in vivo confocal microscopy among study groups.

| **IVCM**<br>**Parameters** | **Controls (n=20)** | **ARSACS (n=8)** | **SPG7 (n=5)** | **p value** |
| --- | --- | --- | --- | --- |
|  |  |  |  | 0.15<sup>a</sup> |
| **CNFD (n/mm<sup>2</sup>)** | 31.2 ± 8.8 | 21.9 ± 4.4 | 15.6 ± 4.4 | 0.16<sup>b</sup> |
|  |  |  |  | 0.10<sup>c</sup> |
|  |  |  |  | **0.045<sup>a</sup>** |
| **CNBD (n/mm<sup>2</sup>)** | 40.6 ± 22.1 | 28.12 ± 4.4 | 18.7 ± 8.8 | **0.01<sup>b</sup>** |
|  |  |  |  | 0.82<sup>c</sup> |
|  |  |  |  | 0.17<sup>a</sup> |
| **CNFL (mm/mm<sup>2</sup>)** | 17.5 ± 2.9 | 15.5 ± 4 | 10.2 ± 0.7 | 0.18<sup>b</sup> |
|  |  |  |  | 0.12<sup>c</sup> |
|  |  |  |  | **0.042<sup>a</sup>** |
| **CTBD (n/mm<sup>2</sup>)** | 59.4 ± 4.4 | 31.2 ± 3 | 28.12 ± 4.4 | **0.025<sup>b</sup>** |
|  |  |  |  | 0.37<sup>c</sup> |
|  |  |  |  | 0.40<sup>a</sup> |
| **CNFA** | 0.008 ± 0.002 | 0.007 ± 0.0017 | 0.005 ± 0.0006 | 0.19<sup>b</sup> |
|  |  |  |  | 0.25<sup>c</sup> |
|  |  |  |  | 0.22<sup>a</sup> |
| **CNFW** | 0.022 ± 0.0015 | 0.021 ± 0.0007 | 0.020 ± 0.0003 | 0.44<sup>b</sup> |
|  |  |  |  | 0.31<sup>c</sup> |
|  |  |  |  | 0.21<sup>a</sup> |
| **CNFrD** | 1.51 ± 0.02 | 1.48 ± 0.04 | 1.44 ± 0.003 | 0.10<sup>b</sup> |
|  |  |  |  | 0.44<sup>c</sup> |

Data are presented as mean ± standard deviation. ARSACS, Autosomal Recessive Spastic Ataxia of Charlevoix-Saguenay; SPG7, Spastic Paraplegia 7; IVCN, In vivo corneal confocal microscopy; CNFD, corneal nerve fiber density; CNBD, corneal nerve branch density; CNFL, corneal nerve fiber length; CTBD, corneal nerve total branch density; CNFA, corneal nerve fiber area; CNFW, corneal nerve fiber width; CNFrD, corneal nerve fractal dimension. a, p value calculated between control and ARSACS; b, p value calculated between control and SPG7; c, p value calculated between ARSACS and SPG7.

Correlations between IVCM parameters and clinical variables are summarized in Table 3. Spearman correlation analysis did not reveal any statistically significant associations between corneal nerve parameters and clinical scale scores, including FARS, SARA, or ADL scale, in either the ARSACS or SPG7 groups. Similarly, no significant correlations were observed between corneal nerve parameters and disease duration in either patient group. Although several moderate correlation coefficients were identified, none reached statistical significance.

**Table 3.** Correlations between in vivo corneal confocal microscopy parameters and clinical features in ARSACS and SPG7 patients.

|  |  | CNFD | CNBD | CNFL | CTBD | CNFA | CNFW | CNFrD |
| --- | --- | --- | --- | --- | --- | --- | --- | --- |
| **FARS** | **ARSACS** | $\rho$ -0.218 | $\rho$ 0.075 | $\rho$ -0.210 | $\rho$ -0.346 | $\rho$ -0.271 | $\rho$ 0.420 | $\rho$ -0.259 |
|  |  | p 0.60 | p 0.85 | p 0.61 | p 0.40 | p 0.51 | p 0.30 | p 0.53 |
| | **SPG7** | $\rho$ -0.078 | $\rho$ -0.216 | $\rho$ 0.615 | $\rho$ 0.216 | $\rho$ -0.359 | $\rho$ -0.461 | $\rho$ 0.615 |
|  |  | p 0.89 | p 0.72 | p 0.26 | p 0.72 | p 0.55 | p 0.43 | p 0.26 |
| **SARA** | **ARSACS** | $\rho$ -0.263 | $\rho$ -0.329 | $\rho$ -0.191 | $\rho$ -0.229 | $\rho$ -0.500 | $\rho$ -0.071 | $\rho$ -0.395 |
|  |  | p 0.52 | p 0.42 | p 0.64 | p 0.38 | p 0.40 | p 0.86 | p 0.33 |
| | **SPG7** | $\rho$ 0.335 | $\rho$ -0.105 | $\rho$ 0.600 | $\rho$ -0.105 | $\rho$ 0.035 | $\rho$ -0.500 | $\rho$ 0.600 |
|  |  | p 0.58 | p 0.86 | p 0.28 | p 0.86 | p 0.93 | p 0.39 | p 0.28 |
| **ADL** | **ARSACS** | $\rho$ -0.378 | $\rho$ -0.628 | $\rho$ -0.371 | $\rho$ 0.341 | $\rho$ 0.035 | $\rho$ 0.275 | $\rho$ -0.395 |
|  |  | p 0.35 | p 0.09 | p 0.36 | p 0.40 | p 0.93 | p 0.51 | p 0.33 |
| | **SPG7** | $\rho$ 0.375 | $\rho$ -0.210 | $\rho$ 0.600 | $\rho$ -0.105 | $\rho$ -0.420 | $\rho$ -0.500 | $\rho$ 0.600 |
|  |  | p 0.52 | p 0.61 | p 0.28 | p 0.86 | p 0.52 | p 0.39 | p 0.28 |
| **Disease duration** | **ARSACS** | $\rho$ 0.206 | $\rho$ 0.257 | $\rho$ 0.253 | $\rho$ 0.150 | $\rho$ 0.443 | $\rho$ 0.192 | $\rho$ 0.494 |
|  |  | p 0.62 | p 0.53 | p 0.54 | p 0.72 | p 0.28 | p 0.64 | p 0.21 |
| | **SPG7** | $\rho$ -0.057 | $\rho$ -0.189 | $\rho$ -0.205 | $\rho$ -0.189 | $\rho$ -0.718 | $\rho$ 0.359 | $\rho$ -0.205 |
|  |  | p 0.92 | p 0.76 | p 0.74 | p 0.76 | p 0.17 | p 0.55 | p 0.74 |

ARSACS: Autosomal Recessive Spastic Ataxia of Charlevoix-Saguenay; SPG7: Spastic Paraplegia 7; CNFD: corneal nerve fiber density; CNBD: corneal nerve branch density; CNFL: corneal nerve fiber length; CTBD: corneal nerve total branch density; CNFA: corneal nerve fiber area; CNFW: corneal nerve fiber width; CNFrD: corneal nerve fractal dimension; FARS: The Friedreich's Ataxia Rating Scale; SARA: the Scale for the Assessment and Rating of Ataxia; ADL: Activities of Daily Living Scale; $\rho$ : Spearman's rank correlation coefficient; p: p value .

## Discussion

In this pilot study, we investigated corneal subbasal nerve plexus alterations using in vivo corneal confocal microscopy in patients with ARSACS and SPG7. Our findings demonstrate that both patient groups exhibit quantitative corneal nerve abnormalities compared with healthy controls, with a decrease in corneal nerve branch density and corneal nerve total branch density. To the best of our knowledge, this is the first study to systematically evaluate corneal nerve involvement in ARSACS and SPG7, providing novel evidence that corneal nerve alterations may represent an ocular manifestation of peripheral neurodegeneration in these spastic ataxias.

Peripheral neuropathy is a well-established component of ARSACS [4] and is increasingly recognized in SPG7, despite the latter traditionally being considered a predominantly central motor disorder [8,9]. The significant reduction in branching-related parameters observed in our cohort is consistent with small fiber involvement and may reflect impaired axonal maintenance and regenerative capacity. Corneal nerve branching metrics are regarded as particularly sensitive indicators of early neuropathic change, as they capture alterations in nerve complexity and remodeling rather than gross axonal loss alone. The selective decrease in CNBD and CTBD, in the absence of statistically significant reductions in corneal nerve fiber density or length, suggests that corneal nerve degeneration in these disorders may initially manifest as simplification of branching architecture before more extensive structural loss becomes evident.

Interestingly, although several additional IVCM parameters—including corneal nerve fiber density, length, area, and fractal dimension—demonstrated numerical reductions in ARSACS and SPG7 patients, these differences did not reach statistical significance. This pattern may indicate that early neurodegenerative changes primarily affect nerve branching complexity rather than overall nerve quantity. Alternatively, the absence of statistical significance may be attributable to limited sample size and inter-individual variability in disease severity. Given the heterogeneity of hereditary ataxias, subtle structural changes may require larger cohorts to detect robust group differences.

Previous neuroimaging studies have demonstrated marked white matter involvement in ARSACS, whereas SPG7 patients often show relatively preserved white matter integrity [21]. In this context, the presence of corneal nerve alterations in both groups in our study should be interpreted cautiously. Our findings may suggest that peripheral corneal small fiber changes could develop independently of white matter involvement and may reflect the contribution of other neuronal injury mechanisms. Given the limited sample size and cross-sectional design, these observations should be considered exploratory.

Given the phenotypic overlap between ARSACS and SPG7 highlighted in clinical practice, the identification of objective biomarkers capable of supporting disease characterization remains an important challenge. Although our results did not demonstrate significant differences in IVCM parameters between the two patient groups, the detection of corneal nerve alterations in both conditions suggests that corneal confocal microscopy may contribute complementary information reflecting peripheral small fiber involvement. Rather than serving as a disease-specific discriminator, IVCM may assist in refining phenotypic assessment when interpreted alongside genetic testing, neuroimaging, and neurophysiological findings. Future studies with larger cohorts are required to determine whether subtle quantitative or pattern-based differences in corneal nerve architecture could support differential evaluation between ARSACS and SPG7.

A previous study utilizing IVCM in other hereditary ataxias, particularly Friedreich ataxia, have demonstrated marked corneal nerve loss and have supported the use of corneal confocal microscopy as a surrogate marker of peripheral neurodegeneration [11]. Our findings extend these observations to ARSACS and SPG7, indicating that corneal nerve alterations may represent a shared feature across genetically distinct ataxic disorders. While retinal and optic nerve abnormalities have been extensively documented in ARSACS and SPG7 [6,7], the cornea has remained largely unexplored despite harboring one of the densest sensory nerve networks in the human body. The identification of corneal nerve involvement therefore suggests a more widespread ocular neurodegenerative process and highlights the anterior segment as an additional site of neural pathology.

Another notable observation was the absence of significant correlations between corneal nerve parameters and clinical severity scales, including FARS, SARA, and ADL scores, as well as disease duration. Several explanations may account for this finding. First, corneal nerve alterations may represent an early or relatively independent manifestation of peripheral nerve involvement that does not directly parallel clinical disability, which primarily reflects motor and functional impairment. Second, the small sample size and variability in disease phenotype may have limited statistical power. Longitudinal studies are needed to determine whether corneal nerve changes progress over time and whether they correlate with clinical deterioration or therapeutic response.

From a pathophysiological perspective, reduced corneal nerve branching complexity may reflect impaired axonal transport, mitochondrial dysfunction, or cytoskeletal instability—mechanisms implicated in hereditary spastic ataxias. Because IVCM allows repeated, non-invasive imaging of peripheral sensory fibers, it offers a unique opportunity to monitor dynamic changes in nerve morphology and may serve as a sensitive biomarker for small fiber pathology. Importantly, corneal confocal microscopy may provide complementary information to neuroimaging by selectively capturing peripheral nervous system involvement that may not be fully reflected by central nervous system assessments.

Several limitations of this study should be acknowledged. The relatively small number of participants, particularly in the SPG7 group, limits generalizability and may have contributed to the lack of statistical significance observed in some parameters. The cross-sectional design precludes assessment of temporal changes in corneal nerve morphology or their relationship with disease progression. In addition, patients with nystagmus and ocular surface disorders were excluded due to technical limitations of image acquisition, which may have introduced selection bias toward less severely affected individuals. Finally, electrophysiological measures of peripheral neuropathy were not systematically correlated with IVCM findings, limiting direct validation against established neurophysiological assessments.

## Conclusion

Our findings demonstrate that corneal subbasal nerve plexus alterations are present in ARSACS and SPG7, with significant reductions in branching-related parameters compared with healthy controls. These results broaden current understanding of ocular and peripheral nervous system involvement in hereditary spastic ataxias and highlight the potential of corneal confocal microscopy as a non-invasive biomarker of small fiber neurodegeneration. Future studies incorporating larger cohorts, longitudinal follow-up, and multimodal correlations with neurophysiological and neuroimaging markers are warranted to further define the clinical utility of corneal nerve assessment in these conditions.

## Data Availability

All data produced in the present study are available upon reasonable request to the authors

## Acknowledgments

The authors gratefully acknowledge the services and facilities provided by the Koç University Research Center for Translational Medicine (KUTTAM). ANB acknowledges the support of the Suna and İnan Kıraç Foundation.

## Author Contributions

**Conceptualization:** Özgür Öztop Çakmak, Atay Vural, Nazan Akkaya, Murat Hasanreisoglu

**Methodology:** Umit Yasar Guleser, Nazan Akkaya, Atay Vural

**Formal analysis and investigation:** Umit Yasar Guleser, Cem Kesim, Melisa Zisan Karslioglu, Nazan Akkaya, Özgür Öztop Çakmak, Atay Vural

**Writing - original draft preparation:** Umit Yasar Guleser, Nazan Akkaya

**Writing - review and editing:** All authors

**Resources:** Atay Vural, Ayşe Nazlı Başak, Özgür Öztop Çakmak, Sibel Ertan, Murat Hasanreisoglu

**Supervision:** Murat Hasanreisoglu, Atay Vural.

